# Time-to-Event Genome-Wide Association Study for Incident Cardiovascular Disease in People with Type 2 Diabetes Mellitus

**DOI:** 10.1101/2023.07.25.23293180

**Authors:** Soo Heon Kwak, Ryan B. Hernandez-Cancela, Daniel A DiCorpo, David E. Condon, Jordi Merino, Peitao Wu, Jennifer A Brody, Jie Yao, Xiuqing Guo, Fariba Ahmadizar, Mariah Meyer, Murat Sincan, Josep M. Mercader, Sujin Lee, Jeffrey Haessler, Ha My T. Vy, Zhaotong Lin, Nicole D. Armstrong, Shaopeng Gu, Noah L. Tsao, Leslie A. Lange, Ningyuan Wang, Kerri L. Wiggins, Stella Trompet, Simin Liu, Ruth J.F. Loos, Renae Judy, Philip H. Schroeder, Natalie R. Hasbani, Maxime M. Bos, Alanna C. Morrison, Rebecca D. Jackson, Alexander P. Reiner, JoAnn E. Manson, Ninad S. Chaudhary, Lynn K. Carmichael, Yii-Der Ida Chen, Kent D. Taylor, Mohsen Ghanbari, Joyce van Meurs, Achilleas N Pitsillides, Bruce M. Psaty, Raymond Noordam, Ron Do, Kyong Soo Park, J Wouter Jukema, Maryam Kavousi, Adolfo Correa, Stephen S. Rich, Scott M. Damrauer, Catherine Hajek, Nam H. Cho, Marguerite R. Irvin, James S. Pankow, Girish N. Nadkarni, Robert Sladek, Mark O. Goodarzi, Jose C. Florez, Daniel I. Chasman, Susan R. Heckbert, Charles Kooperberg, Josée Dupuis, Rajeev Malhotra, Paul S. de Vries, Ching-Ti Liu, Jerome I. Rotter, James B. Meigs, the Cohorts for Heart & Aging Research in Genomic Epidemiology (CHARGE) consortium

## Abstract

**BACKGROUND:** Type 2 diabetes mellitus (T2D) confers a two- to three-fold increased risk of cardiovascular disease (CVD). However, the mechanisms underlying increased CVD risk among people with T2D are only partially understood. We hypothesized that a genetic association study among people with T2D at risk for developing incident cardiovascular complications could provide insights into molecular genetic aspects underlying CVD.

**METHODS:** From 16 studies of the Cohorts for Heart & Aging Research in Genomic Epidemiology (CHARGE) Consortium, we conducted a multi-ancestry time-to-event genome-wide association study (GWAS) for incident CVD among people with T2D using Cox proportional hazards models. Incident CVD was defined based on a composite of coronary artery disease (CAD), stroke, and cardiovascular death that occurred at least one year after the diagnosis of T2D. Cohort-level estimated effect sizes were combined using inverse variance weighted fixed effects meta-analysis. We also tested 204 known CAD variants for association with incident CVD among patients with T2D.

**RESULTS:** A total of 49,230 participants with T2D were included in the analyses (31,118 European ancestries and 18,112 non-European ancestries) which consisted of 8,956 incident CVD cases over a range of mean follow-up duration between 3.2 and 33.7 years (event rate 18.2%). We identified three novel, distinct genetic loci for incident CVD among individuals with T2D that reached the threshold for genome-wide significance (*P*<5.0×10^-8^): rs147138607 (intergenic variant between *CACNA1E* and *ZNF648*) with a hazard ratio (HR) 1.23, 95% confidence interval (CI) 1.15 – 1.32, *P*=3.6×10^-9^, rs11444867 (intergenic variant near *HS3ST1*) with HR 1.89, 95% CI 1.52 – 2.35, *P*=9.9×10^-9^, and rs335407 (intergenic variant between *TFB1M* and *NOX3*) HR 1.25, 95% CI 1.16 – 1.35, *P*=1.5×10^-8^. Among 204 known CAD loci, 32 were associated with incident CVD in people with T2D with *P*<0.05, and 5 were significant after Bonferroni correction (*P*<0.00024, 0.05/204). A polygenic score of these 204 variants was significantly associated with incident CVD with HR 1.14 (95% CI 1.12 – 1.16) per 1 standard deviation increase (*P*=1.0×10^-16^).

**CONCLUSIONS:** The data point to novel and known genomic regions associated with incident CVD among individuals with T2D.

**CLINICAL PERSPECTIVE:** *What is new?:*

- We conducted a large-scale multi-ancestry time-to-event GWAS to identify genetic variants associated with CVD among people with T2D.
- Three variants were significantly associated with incident CVD in people with T2D: rs147138607 (intergenic variant between *CACNA1E* and *ZNF648*), rs11444867 (intergenic variant near *HS3ST1*), and rs335407 (intergenic variant between *TFB1M* and *NOX3*).
- A polygenic score composed of known CAD variants identified in the general population was significantly associated with the risk of CVD in people with T2D.

*What are the clinical implications?:*

- There are genetic risk factors specific to T2D that could at least partially explain the excess risk of CVD in people with T2D.
- In addition, we show that people with T2D have enrichment of known CAD association signals which could also explain the excess risk of CVD.

## INTRODUCTION

Type 2 diabetes mellitus (T2D) is a significant risk factor for cardiovascular disease (CVD), leading in a two- to three-fold higher likelihood of developing CVD. CVD is the leading cause of morbidity and mortality in people with diabetes^1, 2^ and previous studies suggest that life expectancy is reduced by up to eight years in people with T2D^3^. Although CVD mortality rates have declined substantially in the general population in recent decades, this improvement has been less substantial in people with T2D^4^. Various modifiable factors have been extensively investigated, with high blood pressure, hypercholesterolemia, smoking, and diabetes being well-validated risk factors of CVD that are used to estimate the 10-year risk of incident events^5^. However, diabetes itself may explain as much as 75 to 90% of the excess risk of coronary disease in people with T2D^6, 7^.

There are distinct clinical characteristics of CVD in people with T2D that result in less favorable outcomes. People with T2D have an earlier onset of extensive CVD that tends to progress more rapidly than among those without T2D. There is a greater number of atherosclerotic vessels involved and longer diseased vessel segments in people with T2D compared to those without^8^, even after accounting for standard CVD risk factors. The mortality risk at 30 days and one year after acute coronary syndrome was higher in people with T2D than those without T2D^9^. These features, as well as excess CVD risk in people with T2D not solely attributable to more aggregated risk factors, suggest there could be additional risk factors for CVD that are specific to T2D.

Other than traditional risk factors, recent genome-wide association studies (GWASs) have identified at least 204 genetic loci associated with CVD in the general population. These studies have been mostly conducted among European ancestry participants,^10^ which calls for ancestry-diverse studies. So far, identified CVD variants implicate pathways involved in cholesterol metabolism, insulin resistance, thrombosis, inflammation, endothelial function, and vascular remodeling^11^. Genetic risk factors of CVD in the general population are mostly thought to be relevant to people with T2D^12^, leading to the suggestion that genetic predisposition to CVD involves additional loci and/or stronger associations in this group. There are shared genetic loci between T2D and CVD, which include chromosome 9p21.3, *IRS1*, *TCF7L2*, *HNF1A*, and *APOE*^13, 14^. In addition, there is a significant genetic correlation between T2D and CVD^15^. Still, genetic risk factors for CVD in people with T2D have not been thoroughly investigated. Most studies have been underpowered given the stringent significance threshold required for a GWAS and were cross-sectional^13, 16, 17^. To identify genetic risk factors for incident CVD specifically in people with T2D, it is crucial to investigate T2D cases longitudinally and in follow-up studies where diabetes clearly precedes CVD.

We hypothesize that individuals with T2D share an increased CVD genetic burden and that a multi-ancestry GWAS for incident CVD among people with T2D could help identify these signals and infer biological mechanisms underlying the increased CVD risk among people with T2D. To test these hypotheses, we performed a time-to-event GWAS of incident CVD in a large, multi-ancestry sample of people with T2D ensuring that the occurrence of T2D preceded any CVD event.

## METHODS

### Study Design and Participating Cohorts

This is a meta-analysis of ancestry-specific cohort-level time-to-event GWAS for incident CVD in people with T2D, majority of which were from the Cohorts for Heart & Aging Research in Genomic Epidemiology (CHARGE) Consortium^18^. We studied 49,230 participants with T2D from 16 cohorts and of multiple ancestries. In the case of multi-ancestry cohorts, participants were grouped into major ancestries, resulting in 28 ancestry-specific cohort subgroups. Detailed information, including study design, study period, and a brief description of the participants of each cohort, is shown in **Table S1**. All human research was conducted according to the Declaration of Helsinki, and each cohort acquired institutional review board approval. Each participant provided written informed consent.

### Definition of T2D

T2D was defined in each cohort by having one or more of the American Diabetes Association criteria^19^ (**Table S2**). T2D was defined as having at least one of the following conditions: fasting blood glucose (FBG) ≥ 126 mg/dL, hemoglobin A1c (HbA1c) ≥ 6.5%, 2-h glucose by 75-g oral glucose tolerance test ≥ 200 mg/dL, physician-diagnosed diabetes, or use of glucose-lowering medications. Participants with known type 1 diabetes mellitus (T1D) or other specific types of diabetes were excluded. To minimize contamination of T1D, we excluded people with age at diabetes diagnosis of diabetes below 40.

### Definition of Cardiovascular Disease

CVD was defined as a composite of 1) coronary artery disease (CAD), 2) cerebrovascular disease, and 3) death from a cardiovascular cause. CAD included myocardial infarction, stable or unstable angina, percutaneous coronary intervention, coronary artery bypass grafting, and other cohort-defined events (**Table S3**). Cerebrovascular disease included ischemic stroke, hemorrhagic stroke, transient ischemic attack, carotid stenting, endarterectomy, and other cohort-defined events. Death from cardiovascular causes included death from myocardial infarction, stroke, unexpected death presumed to be from ischemic CVD, and other cohort-defined events. An incident CVD event was defined as the first CVD event occurring at least one year after T2D diagnosis. Participants with any CVD event prior to diagnosis of T2D or within one year after diagnosis of T2D were excluded from incident CVD analysis. If a participant had multiple CVD events, only the first event was considered. The source of CVD information varied by each cohort, which included a self-report of the participant, doctor’s notes, cohort visit examinations, linkage to primary care registers and secondary care registers, hospital admissions, and mortality data.

### Cox Proportional Hazards Models

We applied Cox proportional hazards modeling for the time-to-event GWAS. Each single nucleotide variation (SNV) or short insertion-deletion was tested for its association with incident CVD considering observation time and adjusting for covariates. Observation time was defined as years between the age at diagnosis of T2D and age at incident CVD for cases or age at last follow-up for control participants who did not experience CVD event. Before running the Cox proportional hazards model, the model which did not include genotype information was evaluated to determine whether it met the proportional hazard assumption using the cox.zph() function in the ‘Survival’ R package^20^. If a variable violated the assumption, this was resolved either by including an interaction term or by stratifying the variables into 3 to 5 subgroups. Time-to-event GWAS was performed in each ancestry-specific cohort subgroup using either ‘GWASTools’ R package^21^ or ‘gwasurvivr’ R package^22^ (**Table S4**). The primary analysis included age at diagnosis of T2D and sex as covariates, and significant principal components (PCs) were used to adjust for population stratification (basic model). In the full model, body mass index (BMI), current smoking status, treatment for hypertension, systolic blood pressure, total cholesterol, and high-density lipoprotein (HDL) cholesterol level, all of which are used in 10-year risk estimation of CVD, were added to the basic model.

### Cohort-level Analysis

Participants were genotyped using GWAS SNV genotyping arrays (**Table S5**). Each cohort had its specific pre-imputation genotype quality control criteria for call rate, minor allele frequency (MAF), and deviation from Hardy-Weinberg equilibrium. Genotype imputation was performed using either TOPMed reference panel (GRCh38), Haplotype Reference Consortium reference panel (GRCh37), 1,000 Genomes phase 3 reference panel (GRCh37), or population-specific reference panel. All the genotypes aligned on the positive strand of the reference genome. SNVs with poor imputation quality were removed from the analysis in each cohort (INFO score < 0.4 or R^2^ score < 0.3). Within each cohort, analysis was performed separately for four major ancestries: African American (AFR), East Asian (EAS), European (EUR), and Hispanic (HIS). Time-to-event GWAS for incident CVD was performed under the additive genetic model. Familial relationships were handled either by excluding related individuals or by using a robust estimate of variance when relationships were known for the family-based cohort. Cohort-level summary statistics were collected for meta-analysis.

### Meta-Analysis

Summary statistics of the cohort level results underwent standard quality control procedures using EasyQC software^23^. First, the genetic coordinates of GRCh38 were converted to GRCh37/hg19 using LiftOver^24^ software. Then variants with unreliably large effect size (β≥10) or large standard error (≥10) were excluded in each cohort. Variants with minor allele count ≤ 6, or variants with a frequency difference > 0.20 compared to the corresponding ancestry in the 1,000 Genomes phase 3 data were also removed. Allele coding and marker names were harmonized. Meta-analysis of cohort-level summary statistics was conducted using an inverse-variance-weighted fixed-effect method as implemented in METAL^25^. Genomic control was applied at the cohort level to control for possible inflation in the type I error due to residual population stratification. Both overall meta-analysis and ancestry-level meta-analysis were performed. A genome-wide significance threshold was set as *P* < 5.0 × 10^-8^.

### Approximate Conditional Analysis

Approximate conditional analysis using summary statistics was carried out using GCTA-Cojo^26^ (version 1.93.2). Variants within a 120 kilobase (Kb) region (+/- 60 Kb) of the lead signal from the meta-analysis were selected to be included in the conditional analysis. Conditional analyses were approximated using ancestry-specific summary statistics from our association models and then meta-analyzed using a fixed-effect, inverse variance weighted approach, excluding lead variants that only occurred in one ancestry. Linkage disequilibrium was estimated using GCTA by providing the 1000-Genomes reference panel using super populations ‘AFR’ to represent our African ancestry, ‘EUR’ to represent our European ancestry, ‘EAS’ to represent our East Asian ancestry, and the union of ‘AMR’, ‘CEU’ and ‘YRI’ populations to represent our Hispanic ancestry individuals. We applied a multiple testing threshold at each locus when considering variants for distinct secondary signals by dividing 0.05 by the number of variants in the 120 Kb region.

### Fine-Mapping of Distinct Association Signals to Identify 95% Credible Sets

To fine-map distinct association signals, we computed credible sets with 95% confidence. For each variant within +/- 90 Kb of a lead variant with a minor allele count greater than 40, i.e., the most associated variants in a region, we computed ancestry-specific Bayes factors in favor of association using estimated allelic effect sizes and standard errors from each available ancestry-specific meta-analysis. For example, a variant present only in AFR and EUR ancestries would have two Bayes factors corresponding to the AFR and EUR ancestry-specific analyses. To find the Bayes factor (∧_j_) for the *j*th variant in a particular ancestry, we considered

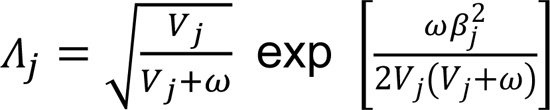

where β_j_ and *V*_j_ represent the effect estimate and variance, respectively, from the ancestry-specific meta-analysis. The constant ω describes the prior variance in allelic effects, which we set to 0.0462.

After computing the Bayes factors, we then calculated the posterior probability that the *j*th variant drives the association signal in a particular ancestry (π_j_) using

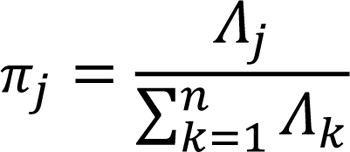

where n denotes the total number of variants within +/- 90 kilobases of the lead variant. This was repeated for all variants in each region.

Finally, ancestry-specific 95% credible sets for each locus were constructed by sorting the variants in a particular locus and ancestry in descending order of probability and finding the smallest subset of the top variants whose sum of probabilities exceeded 0.95.

### Functional Annotations

A genome-wide map of 18 distinct human umbilical vein endothelial cell (HUVEC) chromatin state annotations was retrieved from the Common Metabolic Diseases Genome Atlas (CMDGA, https://cmdga.org/). These chromatin states were characterized from ENCODE^27^ ChIP-seq data using ChromHMM^28^ v1.18. Each variant in each ancestry-specific credible set was matched to its corresponding chromatin state annotation.

### Colocalization of GWAS and eQTL

To estimate the posterior probability of our genome-wide significant variants and eQTL sharing the identical causal variants, we performed a Bayesian colocalization method as implemented in R package ‘coloc’^29^ (cran.r-project.org/web/packages/coloc). eQTL data were obtained from the eQTLGen Consortium^30^ (31,684 whole blood samples), and GWAS variants were extracted from the summary statistics for variants located within one megabase (Mb) of the lead GWAS variants. We defined the variants as colocalized when the posterior probability of a colocalized signal (PP4) was >0.8 as generally recommended.

### Phenome-Wide Association Analysis

For the genome-wide significant variants, we performed additional analyses to gain further insight into how these variants are linked to the pathophysiology of CVD using phenome-wide association analysis. The Common Metabolic Disease Knowledge Portal was used to investigate phenome-wide association (https://hugeamp.org/). As there were 388 phenotypic traits (some of which are interrelated) included in the portal, the significance threshold was conservatively set as *P* < 1.2×10^-4^ (0.05/388).

### Association of known 204 CAD variants

We tested previously reported 204 CAD variants identified in the general population for association with incident CVD in people with T2D^10, 12^. These variants represent genetic risk factors for prevalent CAD in the general population. Adjusting for the multiple comparisons, the association’s significance threshold was set as *P* < 0.00024 (0.05/204). The summary statistics, including effect size (odds ratio) and the *P* value of these variants, were reported previously^12^. In addition, a weighted polygenic score based on these 204 known CAD variants was constructed as previously described^12^ and tested for its association with incident CVD in people with T2D.

## RESULTS

### Study Overview

A total of 49,230 people with T2D who did not have CVD at diagnosis of T2D or within one year of diagnosis were included in the analysis (**Table 1**). There were 16 participating cohorts which were divided into 28 ancestry-specific cohort subgroups (EUR: 14 cohorts, AFR: 8 cohorts, HIS: 3 cohorts, EAS: 3 cohorts). Participants with European ancestry consisted of about 63.2% (N=31,118) of the participants, and the remaining 36.8% (N=18,112) consisted of participants with non-European ancestry (AFR 22.6%, 11,124; HIS 8.8%, 4,325; EAS 5.4%, 2,663). UK Biobank was the largest cohort, representing about 31.8% (N=15,643) of the participants. Among 49,230 participants with T2D, 8,956 developed an incident CVD (event rate 18.2%) over a range of mean follow-up duration between 3.2 and 33.7 years. The CVD event rate by ancestry group was highest in those of African ancestry (25.6%). The detailed clinical characteristics of the participants for each cohort, including mean age at diagnosis of T2D, BMI, smoking status, blood pressure, lipid level, are shown in **Table S6**. In general, and in UK Biobank, those who developed incident CVD had an earlier age at diagnosis of T2D, higher BMI, higher rate of smoking and dyslipidemia (**Table S6**).

**Table 1.**
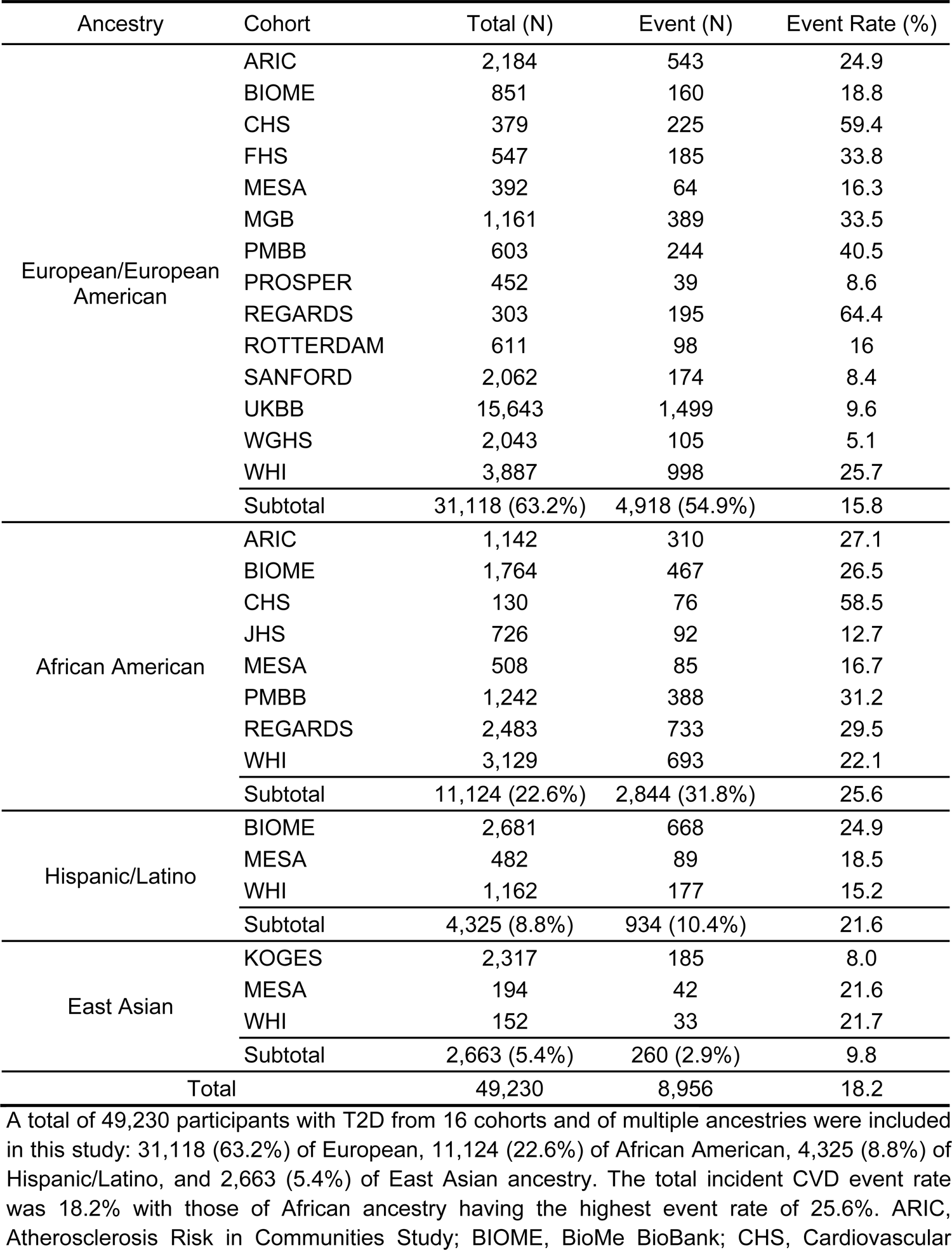
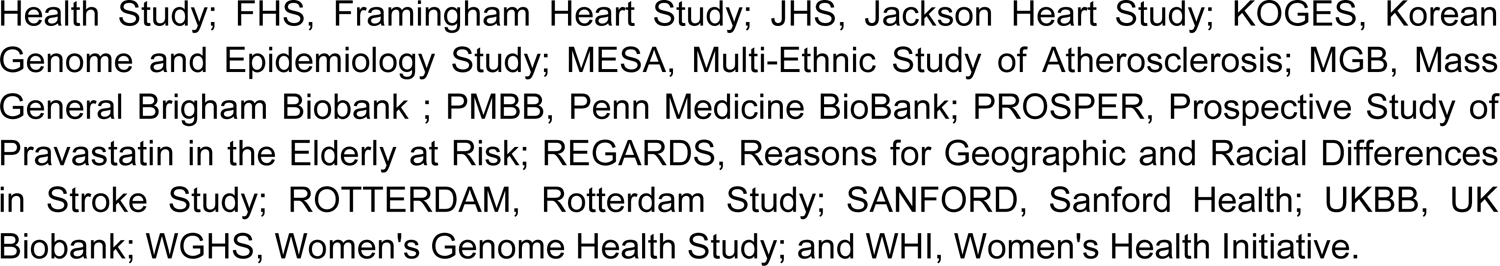
Sample size and event rate according to ancestry and cohort.

| Ancestry | Cohort | Total (N) | Event (N) | Event Rate (%) |
| --- | --- | --- | --- | --- |
| European/European American | ARIC | 2,184 | 543 | 24.9 |
|  | BIOME | 851 | 160 | 18.8 |
|  | CHS | 379 | 225 | 59.4 |
|  | FHS | 547 | 185 | 33.8 |
|  | MESA | 392 | 64 | 16.3 |
|  | MGB | 1,161 | 389 | 33.5 |
|  | PMBB | 603 | 244 | 40.5 |
|  | PROSPER | 452 | 39 | 8.6 |
|  | REGARDS | 303 | 195 | 64.4 |
|  | ROTTERDAM | 611 | 98 | 16 |
|  | SANFORD | 2,062 | 174 | 8.4 |
|  | UKBB | 15,643 | 1,499 | 9.6 |
|  | WGHS | 2,043 | 105 | 5.1 |
|  | WHI | 3,887 | 998 | 25.7 |
|  | Subtotal | 31,118 (63.2%) | 4,918 (54.9%) | 15.8 |
| African American | ARIC | 1,142 | 310 | 27.1 |
|  | BIOME | 1,764 | 467 | 26.5 |
|  | CHS | 130 | 76 | 58.5 |
|  | JHS | 726 | 92 | 12.7 |
|  | MESA | 508 | 85 | 16.7 |
|  | PMBB | 1,242 | 388 | 31.2 |
|  | REGARDS | 2,483 | 733 | 29.5 |
|  | WHI | 3,129 | 693 | 22.1 |
|  | Subtotal | 11,124 (22.6%) | 2,844 (31.8%) | 25.6 |
| Hispanic/Latino | BIOME | 2,681 | 668 | 24.9 |
|  | MESA | 482 | 89 | 18.5 |
|  | WHI | 1,162 | 177 | 15.2 |
|  | Subtotal | 4,325 (8.8%) | 934 (10.4%) | 21.6 |
| East Asian | KOGES | 2,317 | 185 | 8.0 |
|  | MESA | 194 | 42 | 21.6 |
|  | WHI | 152 | 33 | 21.7 |
|  | Subtotal | 2,663 (5.4%) | 260 (2.9%) | 9.8 |
| Total |  | 49,230 | 8,956 | 18.2 |

A total of 49,230 participants with T2D from 16 cohorts and of multiple ancestries were included in this study: 31,118 (63.2%) of European, 11,124 (22.6%) of African American, 4,325 (8.8%) of Hispanic/Latino, and 2,663 (5.4%) of East Asian ancestry. The total incident CVD event rate was 18.2% with those of African ancestry having the highest event rate of 25.6%. ARIC, Atherosclerosis Risk in Communities Study; BIOME, BioMe BioBank; CHS, Cardiovascular
Health Study; FHS, Framingham Heart Study; JHS, Jackson Heart Study; KOGES, Korean Genome and Epidemiology Study; MESA, Multi-Ethnic Study of Atherosclerosis; MGB, Mass General Brigham Biobank ; PMBB, Penn Medicine BioBank; PROSPER, Prospective Study of Pravastatin in the Elderly at Risk; REGARDS, Reasons for Geographic and Racial Differences in Stroke Study; ROTTERDAM, Rotterdam Study; SANFORD, Sanford Health; UKBB, UK Biobank; WGHS, Women's Genome Health Study; and WHI, Women's Health Initiative.

### Loci for Incident CVD in People with T2D

After genotype level quality control, we tested 15,471,776 variants with overall MAF ≥ 1% for association with incident CVD. A plot of expected-by-observed association statistics showed minimal inflation (λ_GC_ = 1.093 for variants with MAF ≥ 1%, λ_GC_ = 1.058 for variants with MAF ≥ 5%) (**Figure 1A**). An association plot of variants with CVD in T2D by chromosomal location identified three variants associated with incident CVD in people with T2D in genome-wide significance (*P* < 5.0×10^-8^) (**Figure 1B**, **Table 2**). The variant rs147138607 (chr1:181855562:G>C, MAF 10.7%) had an HR for incident CVD in T2D of 1.23 (95% CI 1.15 – 1.32, *P* = 3.6×10^-9^) and resides in an intergenic region between the genes *CACNA1E* and *ZNF648* (**Figure 2A**). This variant was most significant in those of African ancestry and was at least nominally significant in the three other ancestry groups (**Table 2**). The second most significant variant rs77142250 (chr4:11444867:T>C, MAF 1.3%) was present at low frequency (1.3%) only in those of African ancestry, had an HR 1.89 (95% CI 1.52 – 2.35, *P* = 9.9×10^-9^), and resides near the gene *HS3ST1* (**Figure 2B**). The third variant rs335407 (chr6:155665441:C>T, MAF 5.5%) had an HR of 1.25 (95% CI 1.16 – 1.35, *P* = 1.58×10^-8^), resides in an intergenic region between the genes *TFB1M* and *NOX3* (**Figure 2C**). This variant was significantly associated with increased risk of CVD in those of European or African ancestry and showed a consistent direction of effects across all ancestry groups. An approximate conditional analysis of these three index variants at each of the three regions of interest showed no evidence of secondary signals (**Figure S1**).

**Figure 1.**
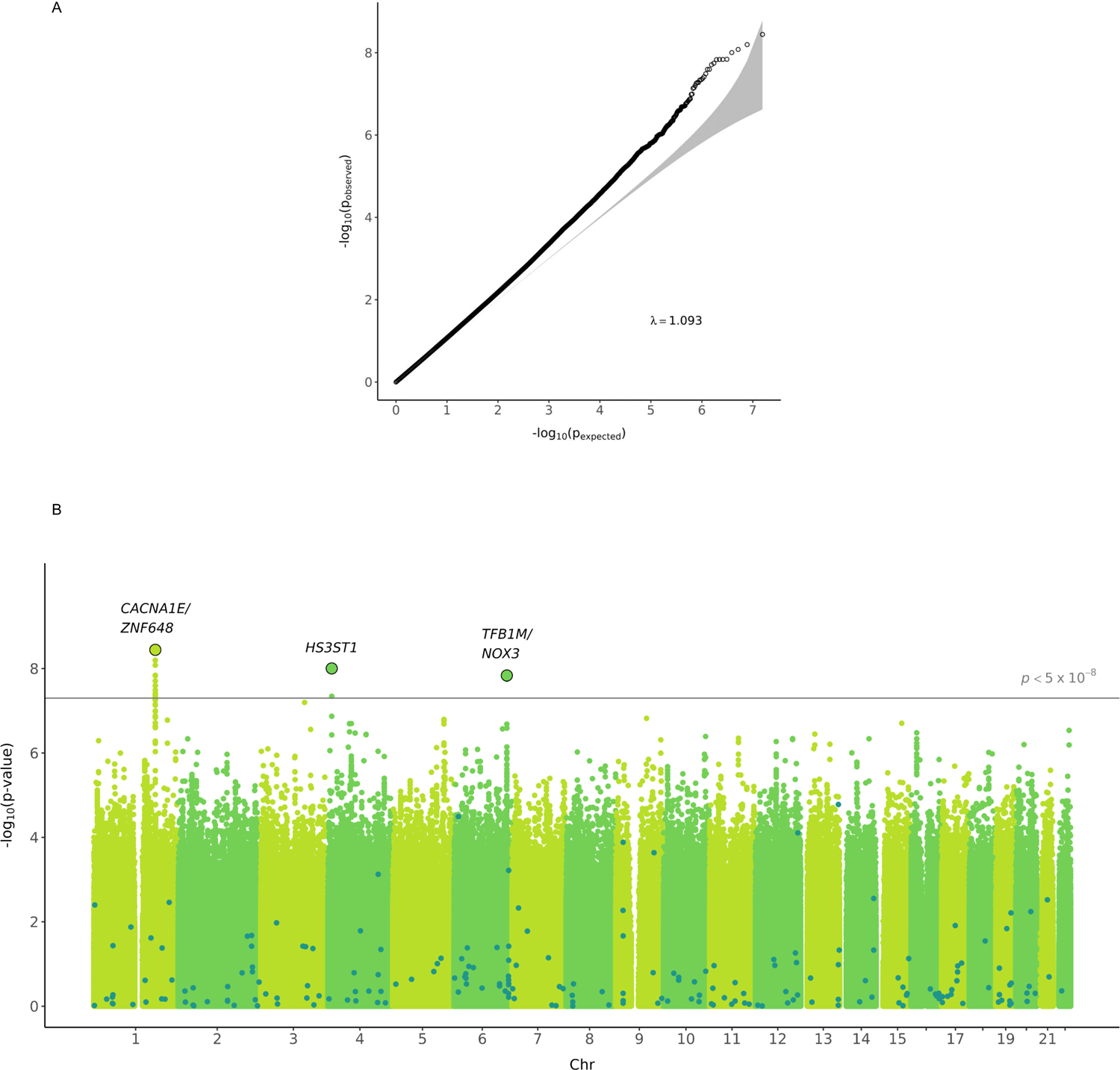
QQ plot and Manhattan plot of the time-to-event GWAS for incident CVD in people with T2D. **A**, QQ plot showing the distribution of the observed *P* values from the meta-analysis of GWAS result against the expected distribution under the null hypothesis. The gray zone indicates the 95% CI. λ_GC_ was 1.093 for variants with MAF ≥ 1% and λ_GC_ was 1.058 for variants with MAF ≥ 5%. **B**, Manhattan plot depicting the significance of all the variants after meta-analysis of GWAS results. SNP locations are plotted on the x-axis according to their chromosomal position. The negative log_10_ of *P* values of time-to-event analysis based on Cox proportional hazard model under the additive model are plotted on the *y*-axis. Dark dots highlight the significance of association for the previously known 204 CAD variants identified in the general population.

**Figure 2.**
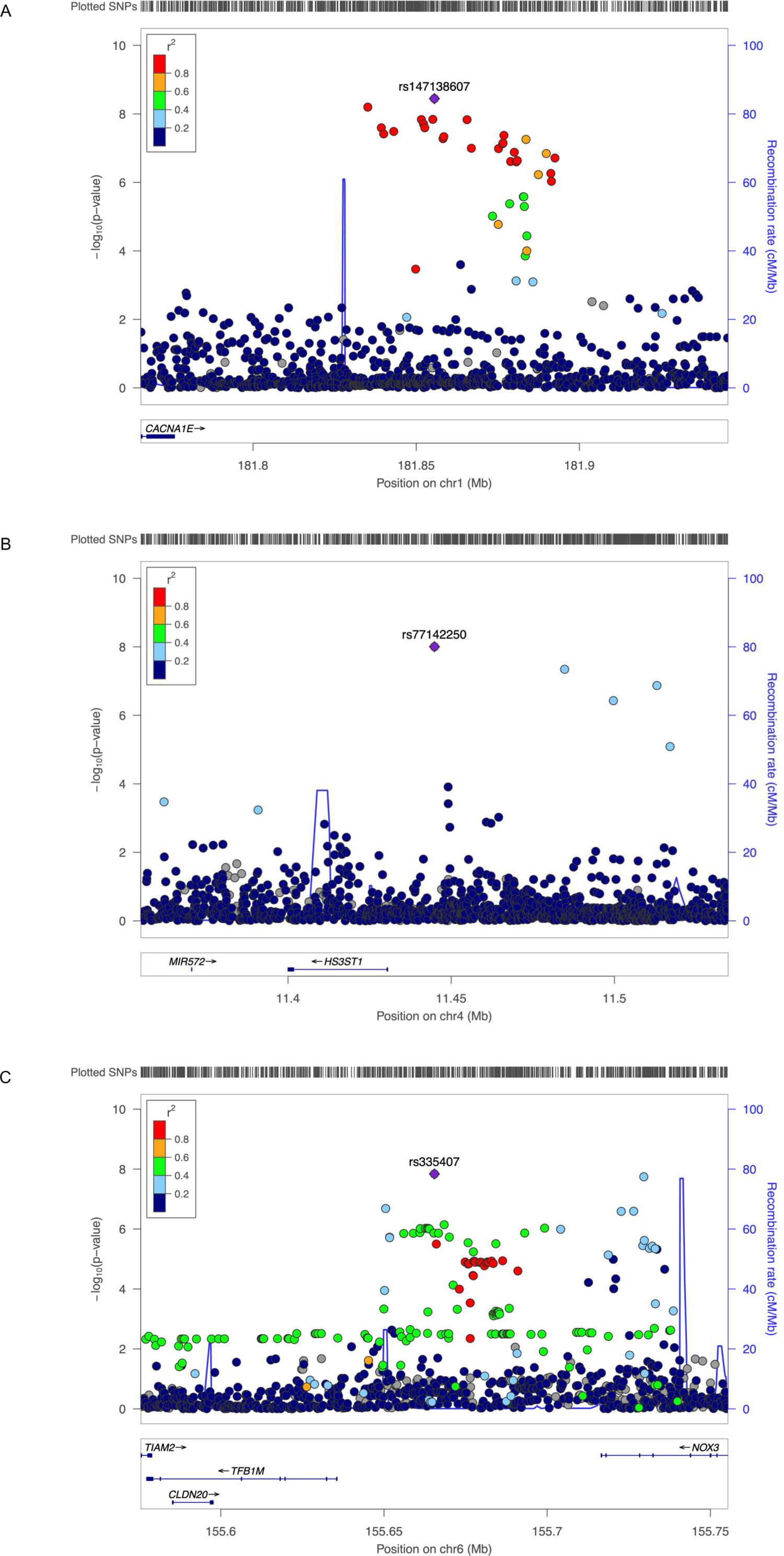
Regional association plots for the three genome-wide significant variants. **A**, rs147138607 near *CACNA1E* and *ZNF648*. **B**, rs77142250 near *HS3ST1*. C, rs335407 near *TFB1M* and *NOX3*. The hash marks above the panel represent the position of each SNP that was genotyped or imputed. The negative log_10_ of *P* values from the Cox regression are shown in the y-axis. Estimated recombination rates are plotted to reflect recombination hot spots. The SNPs in LD with the most significant SNP are color coded to represent their strength of LD based on Europeans for A, and C, and Africans for B.

**Table 2.** Genetic variants significantly associated with incident CVD in people with T2D in basic model.

| CHR | POS (rsID) | Effect Allele | Ancestry | Frequency | HR (95% CI) | *P* | Het *P* | Sample Size |
| --- | --- | --- | --- | --- | --- | --- | --- | --- |
| 1 | 181855562<br>(rs147138607) | G>C | European/European American | 0.018 | 1.20 (1.00-1.44) | 0.047 | 0.756 | 24,457 |
|  |  |  | African American | 0.127 | 1.22 (1.12-1.33) | 2.3x10<sup>-6</sup> | 0.381 | 8,929 |
|  |  |  | Hispanic/Latinx | 0.065 | 1.26 (1.03-1.55) | 0.027 | 0.198 | 3,163 |
|  |  |  | East Asian | 0.050 | 1.55 (1.07-2.25) | 0.021 | 0.469 | 2,511 |
|  |  |  | Combined | 0.107 | 1.23 (1.15-1.32) | 3.6x10<sup>-9</sup> | 0.713 | 39,060 |
| 4 | 11444867<br>(rs77142250) | T>C | African American | 0.013 | 1.89 (1.52-2.35) | 9.9x10<sup>-9</sup> | 0.363 | 9,748 |
| 6 | 155665441<br>(rs335407) | C>T | European/European American | 0.027 | 1.33 (1.19-1.50) | 1.4x10<sup>-3</sup> | 0.664 | 29,910 |
|  |  |  | African American | 0.084 | 1.18 (1.05-1.31) | 3.8x10<sup>-3</sup> | 0.891 | 7,765 |
|  |  |  | Hispanic/Latinx | 0.033 | 1.34 (0.99-1.81) | 0.055 | 0.596 | 3,163 |
|  |  |  | East Asian | 0.026 | 0.92 (0.45-1.88) | 0.810 | 0.541 | 2,511 |
|  |  |  | Combined | 0.055 | 1.25 (1.16-1.35) | 1.5x10<sup>-8</sup> | 0.859 | 43,349 |

Three distinct genetic loci increased risk of incident CVD among individuals with T2D with genome-wide significance in time-to-event analysis ( $P < 5.0 \times 10^{-8}$ ). CHR, chromosome; CI, confidence interval; Het *P*, significance of heterogeneity; HR, hazard ratio; POS, position in GRCh37/hg19; rsID, reference SNP id.

In the fully adjusted model, where covariates of cardiovascular risk factors were included, the statistical significance of the above three variants decreased, which was expected because of the reduced sample size. However, the effect size remained similar. We found two additional genome-wide significant variants that were significant in the full model (**Table S7**). One was an insertion/deletion variant, rs140159474 (chr1:181836968:T>TA, MAF 11.5%) which had an HR 1.23 (95% CI 1.14 – 1.33, *P* = 5.0×10^-8^). This variant was in linkage disequilibrium with the rs147138607 variant, which was significant in the basic model. The second variant was rs76919663 (chr12:61625023:A>G) and was present only in those of African ancestry or Hispanic ancestry, had an HR 1.48 (95% CI 1.30 – 1.67, *P* = 9.8×10^-10^). The nearest gene to this variant was *TAFA2*.

### Insights from Downstream Analysis

We performed fine-mapping analysis for each of the three regions to narrow the number of potentially causal variants using credible set analysis^31^. We constructed a total of nine credible sets: four for the region on chromosome 1, one for chromosome 4, and four for chromosome 6. Each credible set accounted for ≥ 95% of the posterior probability of association with T2D in CVD in its corresponding ancestry-specific analysis. The median credible set size (i.e., number of variants included) for the chromosome 1 locus was 468, with a minimum size of 28 (AFR), and a maximum size of 816 (HIS). Likewise, the median size for the chromosome 6 locus was 637, with a minimum size of 42 (EUR), and a maximum size of 867 (AFR). However, the chromosome 4 credible set only contained 4 variants: rs77142250, rs114281229, rs7677123, and rs77129258. Small credible sets like this are favorable for prioritizing variants for functional follow-up.

Functional interrogation for the three loci included chromatin state annotations from HUVEC cell lines, a system relevant to endothelial function, atherosclerosis, and CVD (**Figure S2**). The chromosome 4 locus contained narrow bands of transcription and enhancer annotations near *HS3ST1* and wider bands dispersed upstream. One of the four variants in the credible set, rs114281229, falls within a region containing an active enhancer annotation. Additionally, rs77129258 resides in a region of weak transcription. Given their proximity, rs114281229 and rs77129258 may affect expression of *HS3ST1*, but further investigation is required to substantiate this. The chromosome 6 locus contained a wide region of active annotations near *TFB1M* and a few smaller regions near *NOX3,* but the size of each credible set in this region was too large to make meaningful functional prioritizations. The chromosome 1 locus contained mostly inaccessible quiescent annotations, with narrow regions of zinc finger protein (ZNF) annotations dispersed throughout.

We used colocalization with expression QTL data from the eQTLGen Consortium to further characterize the three new loci^29^. We found chromosome 6 rs335407 to be a significant eQTL for *TIAM2* in peripheral leukocytes *P* = 7.71×10^-238^. *TIAM2* is essential for endothelial barrier and cell-cell contact maintenance^32^. However, posterior probabilities from colocalization analysis did not support rs335407 as causal for both incident CVD and expression of *TIAM2*. Using a phenome-wide association analysis, we surveyed other phenotypes associated with the three novel loci by interrogating the Common Metabolic Disease Knowledge Portal. Metabolic phenotypes nominally (*P* < 0.05) associated included, for chromosome 1 rs147138607, small artery occlusion and stroke; for chromosome 4 rs7714225, sleep with oxyhemoglobin saturation under 90% and BMI; and for chromosome 6 rs335407, BMI and snoring (**Table S8**).

### Role of Known CAD Variants in People with T2D

We investigated 204 variants of CAD identified in the general population CAD^10, 12^ for its association with CAD in people with T2D (**Table S9**). Among these variants, we observed nominally significant associations with CAD in people with T2D for 38 variants, which included five that were significant after Bonferroni correction (**Figure 3A**). These include rs9349379 at *PHACTR1* locus, rs2891168 at *CDKN2A/2B* locus, rs111245230 at *SVEP1* locus, rs11057830 at *SCARB1* locus, and rs11838776 at *COL4A1/A2.* Among the nominally significant 38 variants, 35 had consistent dirrection of association for CAD in people with T2D and in the general population (binomial *P*=3.1×10^-8^). For the 204 variants, we further observed consistency in the direction of association for risk of CVD between the general population and people with T2D with Spearman coefficient 0.51, *P*=7.2×10^-15^ (**Figure 3B**). Next, we modeled a polygenic score composed of these 204 CAD variants and used this in a model for incident CVD in T2D (**Table 3**). The CAD polygenic score was associated with increased CVD in people with T2D, with an estimated HR of 1.14 (95% CI 1.12 – 1.16) per 1 SD increase. We showed that the association between the CAD polygenic score and CVD differed by ancestry groups (non-significant in East Asians if European derived summary statistics was used). Overall, one standard deviation increase in polygenic score was associated with a 14% increased risk (HR 1.14, 95% CI 1.12 – 1.16) of CVD in people with T2D.

**Figure 3.**
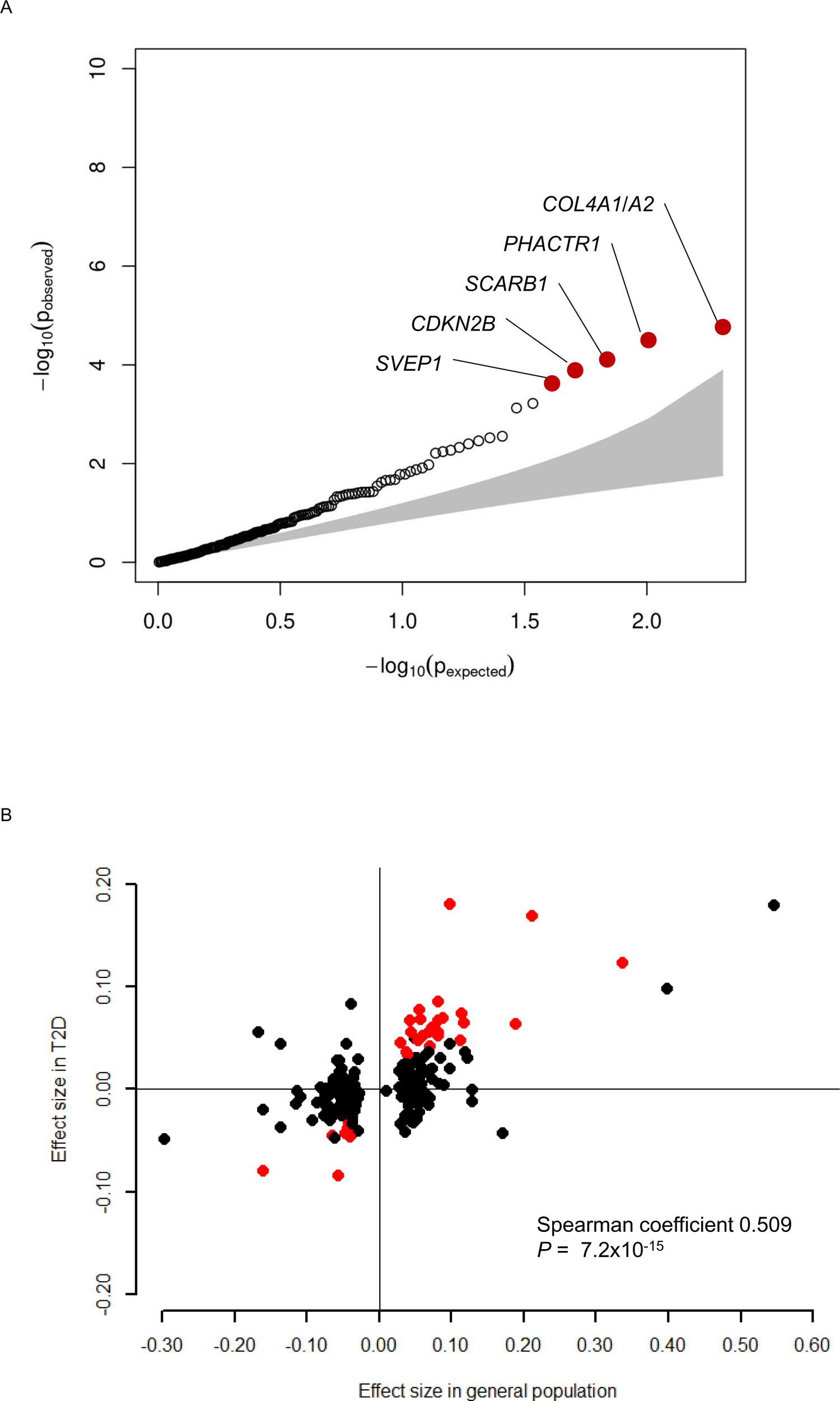
Association of previously identified 204 CAD variants with incident CVD in people with T2D. **A**, QQ plot showing the distribution of the observed *P* values for the 204 CAD variants with risk of incident CVD in people with T2D against the expected distribution under the null hypothesis. The red dots highlight five variants that were significantly associated with incident CVD after Bonferroni correction. **B**, Comparison of the effect size of known 204 CAD variants in the general population and incident CVD in people with T2D. Effect size of the known 204 CAD variants for prevalent CAD in the general population (x-axis, β-coefficient from logistic regression analysis) and incident CVD in people with T2D (y-axis, β-coefficient from Cox regression analysis) are plotted. There was a significant correlation between the effect sizes (Spearman coefficient 0.509, *P*=7.2×10^-15^). The red dots highlight 32 variants that were nominally (*P*<0.05) associated with incident CVD and had same direction of association in the general population.

**Table 3.** Association of polygenic score of 204 known CAD variants and incident CVD in people with T2D.

| Ancestry | HR | 95% CI | Het *P* | *P* |
| --- | --- | --- | --- | --- |
| European/European American | 1.18 | 1.14 - 1.21 | 0.010 | $<1.0 \times 10^{-16}$ |
| African American | 1.10 | 1.05 - 1.15 | 0.255 | $8.3 \times 10^{-5}$ |
| Hispanic/Latinx | 1.10 | 1.03 - 1.18 | 0.474 | 0.0031 |
| East Asian | 0.99 | 0.88 - 1.13 | 0.586 | 0.982 |
| Overall | 1.14 | 1.12 - 1.16 | 0.002 | $<1.0 \times 10^{-16}$ |

Polygenic score of 204 CAD variants discovered from the general population was associated with increased risk of incident CVD in people with T2D. CI, confidence interval; Het *P*, significance of heterogeneity; HR, hazard ratio.

## DISCUSSION

In this study, we sought to identify novel genetic loci associated with incident CVD in people with T2D by performing a time-to-event GWAS. We discovered three distinct variants that reached genome-wide significance: rs147138607 on chromosome 1 between *CACNA1E* and *ZNF648*, rs77142250 on chromosome 4 near *HS3ST1*, and rs335407 on chromosome 6 between *TFB1M* and *NOX3*. All were significant in African ancestry while rs147138607 and rs335407 were also at least nominally significant in European ancestry. None of these variants were significantly associated with CVD in the general population. We found that most CAD variants already known from cross-sectional GWAS in the general population were also associated with incident CVD events in people with T2D, and that a polygenic score composed of 204 CAD variants was associated with incident CVD. To the best of our knowledge, this is the first large-scale genetic association study to investigate genetic risk factors of incident CVD specifically in people with T2D.

The main objective of this study was to identify genetic variants that could explain the excess risk of CVD in people with T2D. We show that people with T2D are enriched with genetic risk factors of CAD observed in the general population: 1) there was an excess number of common single variants known to be associated with CAD in people with T2D, and 2) polygenic score composed of these variants were significantly associated with incident CVD in people with T2D. Furthermore, we identified genetic loci associated with incident CVD, specifically in people with T2D. These variants were not identified as genetic risk factors of CVD in the general population. Taken together, we show that the excess CVD risk in people with T2D is conferred at least in part by excess of known CAD variants and variants that specifically exert their effect in T2D.

The locus on chromosome 1 contained narrow regions of ZNF annotations in a largely quiescent region of inaccessible chromatin. This region has previously been shown to contain a SNP (rs10911021, 226 Kb distal to rs181855562 with R^2^ 0.015) with significant gene-by-diabetes synergism on CVD risk^33^ and all-cause mortality in people with T2D^34^. Located between *ZNF648* and *GLUL*, the rs10911021 variant has been characterized by decreased expression of the *GLUL* gene in human endothelial cells and lower pyroglutamic-to-glutamic acid ratio compared to the protective allele homozygotes, suggesting a mechanistic link between glutamic acid metabolism and CVD risk among people with diabetes^35^. By contrast, not much is known about the role of *ZNF648* itself on T2D and CVD risk, although it has been shown to be a highly conserved gene across species and is essential for erythroid and megakaryocyte differentiation^36^. Further studies are needed to provide mechanistic insights on the potential link between this locus and CVD risk in T2D.

On the chromosome 4 locus, we identified two variants (rs114281229 and rs77129258) in 95% credible set that may play a role in the expression of *HS3ST1* (Heparan Sulfate-Glucosamine 3-Sulfotransferase 1), a sulfated glycosaminoglycan involved in mediating the activities of leukocytes during inflammation through cellular differentiation, proliferation, and homeostasis^37^. *HS3ST1* plays a significant role in the insulin secretory pathway through its effects on membrane depolarization of pancreatic β-cells^38^. An intronic SNP of *HS3ST1* (rs16881446) has previously been implicated in CAD severity and CV events in a candidate gene analysis^37^. One of the variants included in the credible set in our study, rs114281229, fell within a region containing an active enhancer annotation, while the other variant, rs77129258, fell within a region of weak transcription. These findings increase the likelihood that these variants play an important role in the expression of *HS3ST1*, which would need to be verified with functional assays and *in vivo* models of T2D.

Although the size of the credible sets near the *TFB1M* and *NOX3* genes on chromosome 6 was too large to identify specific variants or make functional prioritizations, the rs335407 SNP identified in our study is a known significant eQTL for the *TIAM2* gene in peripheral leukocytes^30^. *TIAM2* (T-cell lymphoma invasion and metastasis 2) is a RAC1 guanine nucleotide exchange factor essential for endothelial barrier and cell-cell contact maintenance^32^. Given its importance in maintaining endothelial cell integrity, *TIAM2* may play an important role in mediating the effects of CVD risk in T2D. Additional studies are needed to validate these findings.

The strengths of this study include the use of time-to-event GWAS for incident CVD rather than performing conventional case-control analysis. This allows us to dissect the temporal relationship between T2D and CVD and assures that T2D qualifies as an exposure variable for CVD and capture CVD events that occur specifically after T2D diagnosis. As far as we are aware of, we have gathered the largest number of T2D samples (N=49,230) with information on incident CVD events (overall event rate 18.2%, N=8,956). We leveraged large-scale biobanks and used age at diagnosis of T2D and CVD event to construct Cox proportional hazards models with observation time defined as years between age at diagnosis of T2D and CVD or last follow-up. This study also benefits from the fact that we included samples from different ancestries and performed a multi-ancestry meta-analysis. As many as 36.8% of the participants were from non-European ancestry. Multi-ancestry meta-analysis is known to increase power where association signal is shared across ancestry groups and improves fine-mapping resolution^39^.

There are certain limitations in this study. First, although we compiled a large number of T2D cases with information on incident CVD, the sample size was still modest. We had limited statistical power compared to the latest GWAS with a case-control design that included more than one million participants^40^. Beyond the three robust association signals we identified, there are likely to be variants lying below the significance threshold. The summary results of our study, which are publicly available on the Common Metabolic Disease Knowledge Portal (https://hugeamp.org), provide a useful resource for further validation and replication. Second, we excluded participants having CVD prior to or within one year of T2D diagnosis. Participants with early onset CVD might have been excluded, and the role of genetic risk factors of CVD discovered in the general population would have been underestimated. However, our focus was to identify genetic risk factors that are specific in people with T2D which could explain the excess risk of CVD, which is the main cause of morbidity and mortality. Finally, there were limitations in performing downstream analysis with our multi-ancestry GWAS results. Most of the publicly available resources, such as GTEx, were developed from the genetic information from European samples, and many of the low-frequency variants in African Americans were absent. In addition, statistical methods for fine mapping, eQTL analysis, and colocalization were not optimized to account for the different LD patterns in multi-ancestry analysis.

In conclusion, we conducted a time-to-event GWAS to identify genetic risk factors of CVD in people with T2D using multi-ancestry cohorts and biobanks. We discovered three loci that are robustly associated with incident CVD and show that known CAD variants identified in the general population are also enriched in people with T2D. These genetic findings might partially explain the excess risk of CVD in people with T2D. Even though the three loci warrant further replication and validation, they point to novel targets for early prevention and treatment of CVD in people with T2D.

## Data Availability

Gentoype data and phenotypes for the participating cohorts of the CHARGE consortium are available via the database of Genotypes and Phenotypes (dbGAP) and the corresponding author upon reasonable request. Summary statistics of the study results will be available on the Common Metabolic Disease Knowledge Portal (https://hugeamp.org).

